# Primary Care Obesity Management at the Threshold of the GLP-1 Era: A Survey-Based Change Readiness Assessment

**DOI:** 10.64898/2026.04.01.26349998

**Authors:** Mary Ales, Christopher Larrison, Shelly Rodrigues

## Abstract

**Background:** Between 2021 and 2022, primary care obesity management was entering the early diffusion phase of newer anti-obesity pharmacotherapy, as GLP-1–based treatments began reshaping expectations. However, it was unclear whether primary care clinicians and practice environments were prepared to deliver comprehensive obesity care. (1,2)

**Methods:** In 2021–2022, we surveyed 276 clinicians from three cohorts: an opt-in national physician panel (Cohort A), clinicians from an integrated health system (Cohort B), and clinicians from a rural accountable care organization (Cohort C). The survey, informed by formative patient and physician focus groups conducted in 2021, assessed current and desired competence, attitudes, confidence, perceived forces for change, and barriers to obesity care. Analyses were descriptive (means and standard deviations).

**Results:** Across cohorts, desired competence exceeded current competence. The largest gaps involved recommending behavioral interventions, developing comprehensive care plans, and providing ongoing obesity management support. Attitudes toward obesity care were generally favorable, while confidence that current practices reflected best practice was only moderate. Professional and personal forces for change were moderate, patient-driven motivators were moderate to high, whereas social (peer/organizational) reinforcement was weak. Reported barriers extended beyond knowledge deficits to include patient engagement, competing demands, cost, and practical constraints.

**Conclusions:** At the threshold of the GLP-1 era, primary care clinicians were motivated to improve obesity care but lacked consistent support to deliver comprehensive management. The relative absence of peer and organizational reinforcement suggests that readiness for change reflected not only individual knowledge and attitudes, but also the degree of peer and organizational reinforcement that supports comprehensive obesity care in routine practice.

## Introduction

By 2021–2022, primary care obesity management was entering a period of rapid change. Newer pharmacologic options were beginning to reshape the treatment landscape (1), yet most primary care clinicians were practicing in settings marked by limited time, patient perceived stigma, uneven training, and uncertain system support (2). This transitional period offers an important view of obesity care just before the GLP-1 era became fully established. Contemporary guidance now supports anti-obesity pharmacotherapy, as part of comprehensive management for appropriate patients (3–5).

This study examined readiness for change in obesity care, not simply knowledge deficits. Drawing on Fox’s framework describing physician change as shaped by personal, professional, and social forces, we focused on how clinicians perceived their current and desired competence, the barriers they faced, and the forces shaping practice change (6,7).

Our central question was: what did readiness for change in primary care obesity management look like in 2021–2022? We focused on (1) where clinicians perceived the largest competence gaps, and (2) what forces and barriers shaped the feasibility of improvement.

## Methods

### Study design and survey development

This was a survey-based, multi-cohort needs assessment. During instrument development, two patient focus groups and two physician focus groups were conducted in spring 2021 to identify the competencies and practice issues most relevant to obesity care in the primary care setting.

Using findings from the formative focus groups, project discussions, expert input, and the conceptual distinction among personal, professional, and social forces for change, the team developed a change-readiness inventory focused on primary care obesity management. The instrument assessed self-reported current and desired competence, attitudes, confidence, perceived forces for change, and barriers to obesity care. Competencies reflected patient-centered obesity management and included assessment, shared decision-making, care planning, longitudinal care, lifestyle counseling, and pharmacologic treatment.

### Survey sample

The survey dataset used in this manuscript includes 276 primary care clinicians across three cohorts. Cohort A included 202 physicians recruited through national panel provider. Cohort B included 46 clinicians from an integrated health system, and Cohort C included 28 clinicians from a rural accountable care organization. (Table 1).

**Table 1.**

|  | Cohort A | Cohort B | Cohort C | Total |
| --- | --- | --- | --- | --- |
| **Profession** |  |  |  |  |
| Physician | 202 (100.0%) | 33 (71.7%) | 15 (53.6%) | 250 (90.6%) |
| Nurse Practitioner | 0 (0.0%) | 12 (26.1%) | 11 (39.3%) | 23 (8.3%) |
| Physician Assistant | 0 (0.0%) | 1 (2.2%) | 2 (7.1%) | 3 (1.1%) |
| **Specialty** |  |  |  |  |
| Family Medicine | 101 (50.0%) | 38 (82.6%) | 24 (85.7%) | 163 (59.1%) |
| Internal Medicine | 101 (50.0%) | 8 (17.4%) | 2 (7.1%) | 111 (40.2%) |
| Ob/Gyn | 0 (0.0%) | 0 (0.0%) | 2 (7.1%) | 2 (0.7%) |
| Geriatrics | 0 (0.0%) | 0 (0.0%) | 1 (3.6%) | 1 (0.4%) |
| **Years in practice** |  |  |  |  |
| 0–5 years | 5 (2.5%) | 17 (37.0%) | 14 (50.0%) | 36 (13.0%) |
| 6–10 years | 13 (6.4%) | 4 (8.7%) | 3 (10.7%) | 20 (7.2%) |
| 11–15 years | 25 (12.4%) | 8 (17.4%) | 0 (0.0%) | 33 (12.0%) |
| 16–20 years | 39 (19.3%) | 4 (8.7%) | 4 (14.3%) | 47 (17.0%) |
| 20 years plus | 120 (59.4%) | 13 (28.3%) | 7 (25.0%) | 140 (50.7%) |

Values are n (%).

### Measures and analysis

Competence items asked respondents to rate their current and desired agreement with competency statements on 5-point Likert-type scales. Gap scores were defined as desired competence minus current competence (Table 2). Additional items assessed attitudes toward obesity care, confidence that current practice reflected best practice, perceived forces for change, and barriers to change using similar response scales. Current and desired competence item-level results are provided in Supplemental Tables S1 and S2.

**Competency Table 2.**

|  | Gap A | Gap B | Gap C |
| --- | --- | --- | --- |
| Assess a patients' readiness to engage in weight management. | 0.75 (1.06) | 1.04 (1.01) | 1.11 (0.83) |
| Communicate the benefits to overall health of sustained weight loss to persons with obesity (PwO). | 0.45 (0.94) | 0.91 (1.07) | 1.04 (0.79) |
| Develop a comprehensive, personalized obesity management care plan taking into account the patient's unique circumstances | 1.08 (1.25) | 1.52 (1.01) | 1.54 (1.04) |
| Elicit a comprehensive obesity-focused medical history. (Weight trends, previous weight loss attempts, etc.) | 0.79 (1.12) | 1.07 (0.98) | 1.32 (1.02) |
| Engage the patients in shared decision-making by incorporating their values and preferences. | 0.69 (1.05) | 0.91 (1.05) | 1.00 (0.94) |
| Perform and document a comprehensive physical examination for the assessment of obesity and the patient's overall health. | 0.35 (0.92) | 0.74 (0.95) | 0.71 (0.81) |
| Provide on-going care including the assessment of patient progress (weight, energy, function, effect on obesity related comorbidities) and supporting maintenance. | 0.95 (1.12) | 1.24 (1.04) | 1.14 (0.85) |
| Recommend (or refer for) personal and achievable behavioral interventions | 1.10 (1.11) | 1.63 (1.08) | 1.71 (1.05) |
| Recommend (or refer for) personal and achievable nutrition interventions | 0.89 (1.06) | 1.24 (1.02) | 1.25 (0.97) |
| Recommend (or refer for) personal and achievable physical activity interventions | 0.78 (1.12) | 1.22 (1.05) | 1.36 (0.87) |
| Refer appropriate patients for surgical treatments | 0.57 (1.08) | 0.93 (0.95) | 1.07 (1.30) |
| Utilize pharmacological treatments when indicated | 0.77 (1.08) | 1.33 (0.99) | 1.04 (1.20) |

Values are mean (SD). Likert scale: 1 = low agreement; 5 = high agreement. Gap = desired competence – current competence.

Survey responses were analyzed descriptively. Mean scores and standard deviations were calculated for scaled items, and respondent characteristics were summarized using counts and percentages. Gap scores were used to identify differences between current and desired competence across domains. Because the cohorts differed in sampling approach and composition, findings were interpreted descriptively rather than as formal statistical comparisons across cohorts.

## Results

### Sample characteristics

The survey dataset included 276 primary care clinicians across three cohorts. Cohort A included 202 physicians recruited through an opt-in research firm, Cohort B included 46 clinicians from an integrated health system, and Cohort C included 28 clinicians from a rural accountable care organization. Across all respondents, 250 were physicians, 23 were nurse practitioners, and 3 were physician assistants. Cohort A had a more experienced workforce, with 59.4% reporting more than 20 years in practice, compared with 28.3% in Cohort B and 25.0% in Cohort C. Clinicians with 0–5 years in practice were more common in Cohort B (37.0%) and Cohort C (50.0%) than in Cohort A (2.5%). Additional participant characteristics are presented in Table 1.

### Current competence

Across cohorts, current competence scores were generally higher in assessment and communication domains and lower in domains requiring longitudinal or personalized care. For example, mean current competence for performing and documenting a comprehensive physical examination ranged from 3.67 to 4.11, whereas competence for developing a comprehensive personalized care plan ranged from 2.89 to 3.29 and competence for recommending or referring for achievable behavioral interventions ranged from 2.93 to 3.32 (Supplemental Table S1).

### Desired competence and competence gaps

Desired competence scores were high across cohorts, and desired competence consistently exceeded current competence. The largest gaps clustered in domains central to longitudinal, individualized obesity care rather than basic assessment tasks (**Table 2; Figure 1; Supplemental Table S2**). The highest gaps involved recommending achievable behavioral interventions and developing comprehensive, personalized care plans, followed by needs related to ongoing management. Cohorts B and C generally reported larger gaps than Cohort A across the highest-ranked competencies. Attitudes and confidence

**Figure 1.**
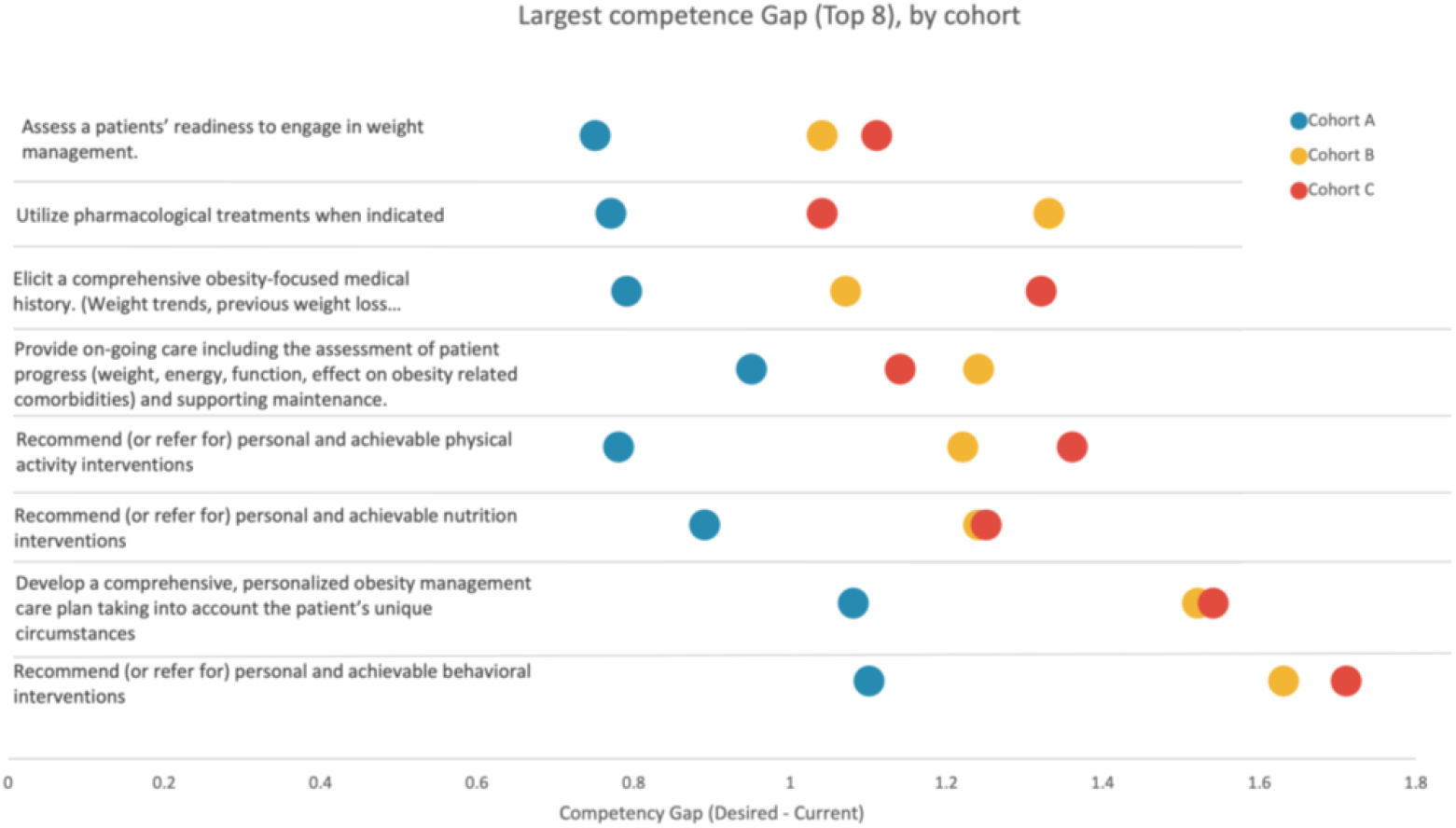
Largest competence gaps in primary care obesity management, by cohort (Top 8 items). Mean competence gap. Desired competence minus current competence. Five-point scale for the eight competencies with the highest overall gaps across three cohorts. Points show cohort-specific mean gaps. Larger values indicate greater perceived need for improvement.

Attitudes toward obesity care were generally favorable across all cohorts. Respondents strongly endorsed the statement that obesity is a chronic disease, with scores ranging from 4.57 to 4.71. They also generally agreed that they could make a difference for patients with obesity, with scores ranging from 3.88 to 4.18. Confidence that current practices reflected best practice in obesity management was more moderate, with scores ranging from 2.98 to 3.50 (Table 3).

**Table 3.**

| Attitude | Cohort A | Cohort B | Cohort C |
| --- | --- | --- | --- |
| I believe I can make a difference for my patients with obesity. | 3.88 (0.89) | 3.91 (1.05) | 4.18 (1.12) |
| I believe that obesity is a chronic disease. | 4.57 (0.62) | 4.70 (0.51) | 4.71 (0.81) |
| I believe that obesity is an addiction. | 3.42 (1.08) | 3.02 (1.24) | 3.79 (0.99) |
| My current practices related to obesity management are acceptable to me. | 3.32 (1.02) | 2.89 (1.08) | 2.96 (1.07) |
| I need to make changes related to the way I treat obesity. | 3.45 (1.04) | 3.33 (1.19) | 3.46 (1.07) |
| **Confidence** |  |  |  |
| How confident are you that your current practices reflect best practices related to obesity management? | 3.50 (0.92) | 2.98 (1.06) | 3.18 (0.90) |

Values are mean (SD). Likert scale: 1 = low; 5 = high.

### Forces for change

Across cohorts, professional and personal forces for change were generally moderate (Table 4; Figure 2). Clinicians reported a moderate desire to be more knowledgeable about obesity care (3.49–3.89) and moderate influence of personal experience with obesity among family and friends (3.42–4.21). Patient-driven motivators were moderate to high, particularly perceived requests from patients for more assistance (3.25–3.91). In contrast, social (Fox) forces were consistently lower, including perceived pressure from colleagues (1.54–2.13) and perceived system encouragement to change obesity care practices (2.19–2.91).

**Table 4.**
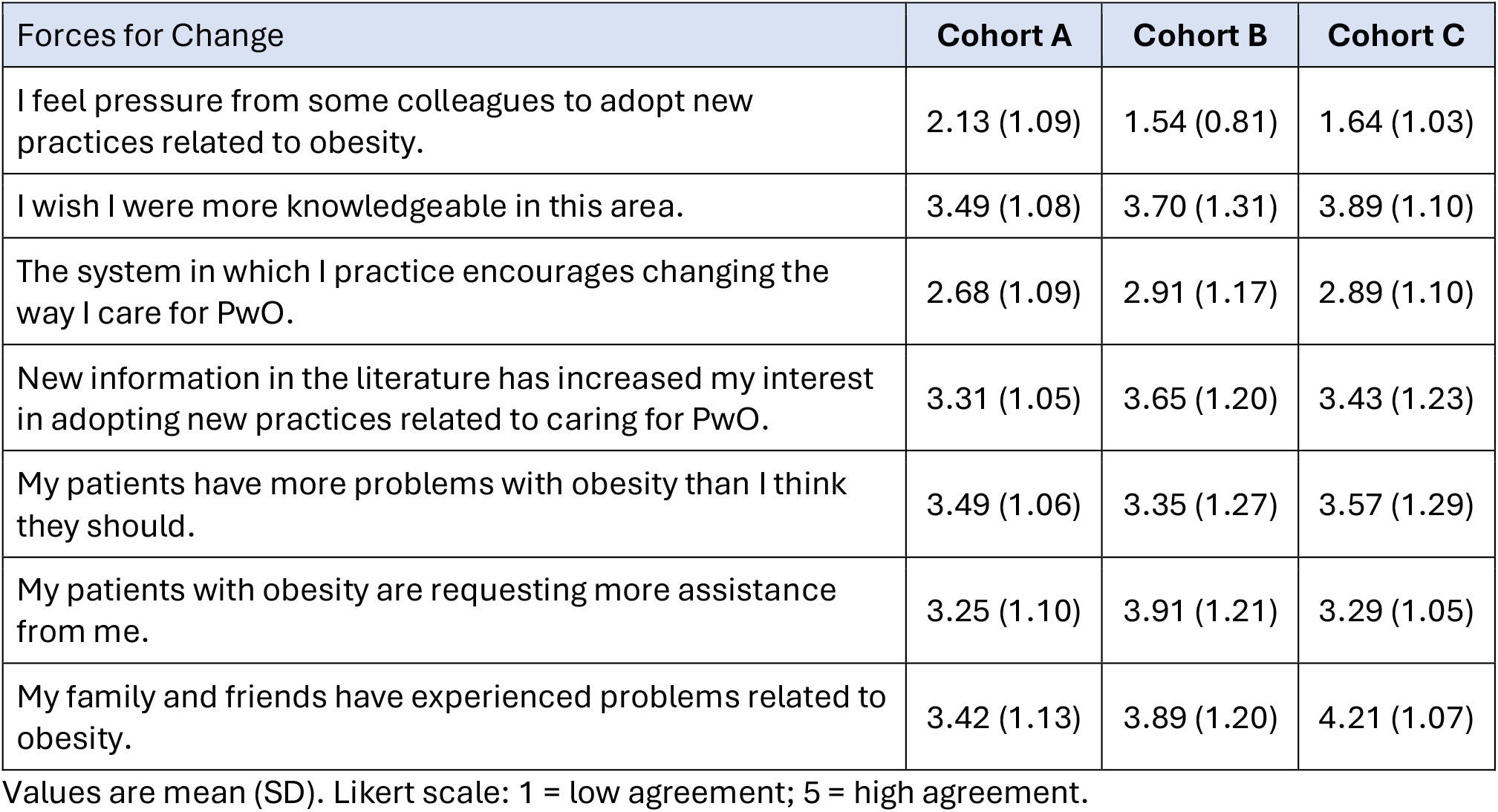

**Figure 2.**
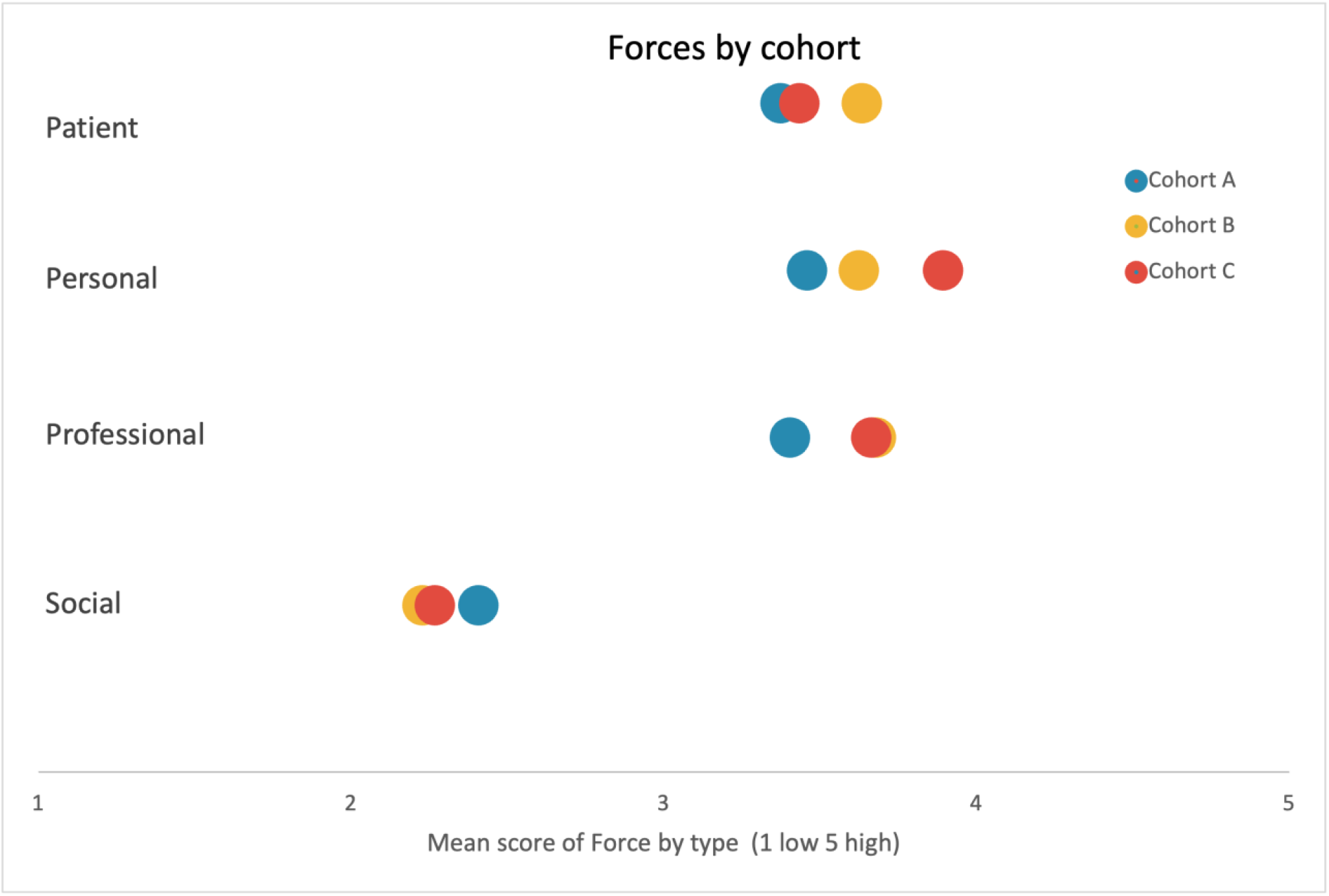
Forces for change (Fox domains + patient-driven), by cohort. Mean agreement (1–5) for forces that may influence change in obesity care, grouped into Fox’s professional, personal, and social domains, with patient-driven factors shown separately

### Barriers to change

Respondents reported barriers across multiple domains, but no single barrier stood out as overwhelmingly high or negligible across cohorts. For example, patient reluctance to engage in a comprehensive weight management plan ranged from 3.45 to 3.46, while discomfort with prescribing current weight-loss medications ranged from 2.32 to 2.94 (Table 5).

**Table 5.**

| Barriers | Cohort A | Cohort B | Cohort C |
| --- | --- | --- | --- |
| I am not comfortable prescribing the current weight loss medications. | 2.94 (1.31) | 2.37 (1.31) | 2.32 (1.22) |
| I do not have access to weight loss specialists (including obesity medicine physicians, nutritionists, counseling, exercise, bariatric surgeons, etc.) | 2.39 (1.20) | 2.22 (1.32) | 2.54 (1.32) |
| My patients are reluctant to engage in a comprehensive weight management plan. | 3.45 (0.92) | 3.59 (1.20) | 3.46 (1.04) |
| My practice team does not embrace changing our approach to obesity. | 2.61 (1.08) | 1.67 (0.79) | 2.00 (0.86) |
| Other responsibilities prevent me from engaging with my patients in their weight loss journey. | 3.15 (1.14) | 3.35 (1.42) | 3.14 (0.97) |
| Reimbursement/payment is insufficient for me to make changes in treatment of PwO. | 3.08 (1.15) | 2.78 (1.44) | 3.00 (1.33) |
| Societal acceptance of obesity makes treatment difficult. | 3.41 (1.08) | 2.89 (1.22) | 3.39 (1.17) |
| The cost of treatment is prohibitive. | 3.30 (1.12) | 3.98 (1.24) | 3.71 (0.98) |
| Understanding and addressing cultural influences on a patient's lifestyle is difficult. | 3.32 (1.03) | 2.98 (1.13) | 2.89 (0.79) |

Values are mean (SD). Likert scale: 1 = low agreement; 5 = high agreement.

## Discussion

This study captured primary care obesity care at a transitional moment when newer pharmacologic options were beginning to reshape treatment. Across cohorts, clinicians generally viewed obesity as a chronic disease, wanted greater competence, and expressed motivation to improve care. Less developed was the practice environment needed to support change.

The largest gaps centered on behavioral intervention, personalized care planning, and ongoing longitudinal management. These are core elements of chronic disease care. The pattern suggests that the challenge was less about recognizing obesity as important and more about translating that recognition into comprehensive care over time.

The gap profile in **Figure 1** suggests that educational needs in 2021–2022 were not primarily concentrated in foundational obesity assessment or in recognizing obesity as a chronic disease. Instead, the largest gaps reflected the “middle and late” work of comprehensive care: translating patient goals into feasible plans, supporting behavioral change through achievable interventions or referrals, and sustaining longitudinal follow-up. From an educational design perspective, these findings point toward curricula that emphasize care planning, team-based counseling/referral pathways, and ongoing management processes—not knowledge acquisition alone.

No single barrier dominated the findings; instead, respondents described a set of moderate constraints across patient, clinical, and practical domains. This suggests that the challenge in obesity care may lie less in overcoming one major obstacle than in building practice environments that reduce the cumulative friction of many smaller ones.

The forces-for-change findings are especially notable. Fox and colleagues described physician change as influenced by personal, professional, and social forces. In this dataset, clinicians reported greater momentum to change from professional and personal factors—including perceived knowledge gaps, new information, patient requests for assistance, and personal experience—than from colleagues or organizational encouragement. This pattern suggests that clinicians may be open to change even when the surrounding practice environment provides relatively weak reinforcement (6,7).

The forces-for-change pattern in **Figure 2** adds an important readiness signal. Clinicians reported meaningful motivation from professional and personal factors and from patient-driven demand, yet they reported comparatively weak reinforcement from social forces—particularly peers and organizational encouragement. This combination suggests a readiness profile in which clinicians may be willing to improve, but may lack the local norms, workflows, and structural supports that make comprehensive obesity care routine. Educational strategies that include practical implementation supports (e.g., workflow tools, role clarity, and longitudinal follow-up structures) may therefore be more aligned with the barriers and forces identified than traditional, clinician-only approaches.

That pattern may reflect a broader shift in the ecology of physician learning and change. Fox’s work emerged in a period when physicians often had greater autonomy to translate new knowledge into practice. Contemporary primary care is more team-based, system-dependent, and shaped by workflows, incentives, and competing demands (8,9). In that setting, individual motivation may remain necessary, but it is less likely to be sufficient. Obesity care may be especially vulnerable because, despite being a chronic disease, it has not historically had the same shared structures that support management of conditions such as diabetes and hypertension.

Descriptive differences across cohorts should be interpreted cautiously. Cohort A, the older physician-only group, reported somewhat smaller gaps and greater confidence, whereas Cohorts B and C reported larger gaps. Because the cohorts differed in profession mix, years in practice, and practice context, these patterns should not only be interpreted simply as setting effects.

Primary care entered this rapidly changing treatment era with meaningful individual readiness but incomplete system readiness. More effective pharmacotherapy did not eliminate the need for communication, care planning, follow-up, team support, or organizational alignment. If anything, it increased the importance of those elements.

These findings also have implications for continuing education. Traditional knowledge-based CME remains necessary, especially in areas such as pathophysiology, treatment options, and pharmacologic therapy. However, education focused only on the individual clinician is unlikely to be sufficient when the most prominent gaps involve care planning, longitudinal management, and implementation within imperfect systems. Educational and implementation strategies should therefore also address stigma-aware communication, shared care planning, longitudinal follow-up, team roles, workflow design, and system-level support.

## Limitations

This study has several limitations. First, the survey reflects self-reported perceptions rather than observed practice or patient outcomes. Second, findings across cohorts are descriptive because the groups differed in profession mix, specialty mix, years in practice, and organizational context; we did not conduct formal statistical comparisons across cohorts. The addition of two practice-based cohorts increased heterogeneity and representation, but also increases caution in interpreting between-cohort differences. Third, formative focus groups informed survey development rather than serving as a formal qualitative component of the study. Fourth, data were collected during 2021–2022, an early diffusion period for newer anti-obesity pharmacotherapy, and therefore capture a transitional moment rather than the current state of obesity care. Finally, the opt-in sampling approach for Cohort A and differences in sampling across cohorts may limit generalizability beyond the participating groups.

## Conclusion

This survey-based needs assessment suggests that primary care entered the early GLP-1 era with substantial clinician motivation but limited professional and organizational reinforcement for change. Readiness was therefore uneven: clinicians generally wanted to improve, but the systems around them were not consistently configured to support comprehensive obesity care.

The key implication is that the problem was not persuading clinicians that obesity matters. Many already believed that. The larger challenge was supporting motivated clinicians work within systems that did not yet strongly operationalize comprehensive obesity care. For medical education, this means pairing clinical knowledge with practical strategies that support implementation, team-based care, and ongoing management.

## Data Availability

All data produced in the present study are available upon reasonable request to the authors

## Acknowledgements

This research was funded by an independent educational grant from Pfizer Learning and Change. We thank the patients and physicians who participated in the formative focus groups and the expert panel that reviewed the findings from this project and offered their insights and perspectives.

## Declarations

### Consent to Publish

Not applicable. The manuscript does not contain any individual person’s data in any form.

### Data Availability

All data not included in the manuscript are available on request.

### Authors’ Contributions

MA, SR and CL led the study design, implementation oversight, and data analysis. MA led manuscript drafting. All authors reviewed and approved the final manuscript.

## Acknowledgements

The authors thank the physicians and patients who participated in the focus groups, the organizations for survey distribution and the numerous physicians and obesity experts who discussed obesity management with us.

## Supplemental Data

### Competency Present Supplemental Data 1

| Competency Present Supplemental Data 1 | Present A | Present B | Present C |
| --- | --- | --- | --- |
| Assess a patients' readiness to engage in weight management. | 3.65 (0.98) | 3.50 (1.13) | 3.64 (0.87) |
| Communicate the benefits to overall health of sustained weight loss to persons with obesity (PwO). | 4.07 (0.87) | 3.72 (1.20) | 3.79 (0.88) |
| Develop a comprehensive, personalized obesity management care plan taking into account the patient's unique circumstances | 3.29 (1.12) | 2.89 (1.16) | 3.11 (0.99) |
| Elicit a comprehensive obesity-focused medical history. (Weight trends, previous weight loss attempts, etc.) | 3.55 (1.09) | 3.24 (1.18) | 3.32 (1.16) |
| Engage the patients in shared decision-making by incorporating their values and preferences. | 3.73 (0.97) | 3.65 (1.16) | 3.79 (0.99) |
| Perform and document a comprehensive physical examination for the assessment of obesity and the patient's overall health. | 4.10 (0.94) | 3.67 (1.16) | 4.11 (0.92) |
| Provide on-going care including the assessment of patient progress (weight, energy, function, effect on obesity related comorbidities) and supporting maintenance. | 3.44 (1.10) | 3.13 (1.13) | 3.61 (0.88) |
| Recommend (or refer for) personal and achievable behavioral interventions | 3.32 (1.08) | 2.93 (1.08) | 2.96 (1.10) |
| Recommend (or refer for) personal and achievable nutrition interventions | 3.58 (1.05) | 3.46 (0.96) | 3.54 (1.00) |
| Recommend (or refer for) personal and achievable physical activity interventions | 3.73 (1.06) | 3.41 (1.02) | 3.39 (1.03) |
| Utilize pharmacological treatments when indicated | 3.34 (1.18) | 3.15 (1.09) | 3.46 (0.96) |
| Refer appropriate patients for surgical treatments | 3.78 (1.02) | 3.74 (0.93) | 3.50 (1.17) |

Values are mean (SD). Likert scale: 1 = low agreement; 5 = high agreement.

### Competency Desired Supplemental Data 2

| Competency Desired Supplemental Data 2 | Desired A | Desired B | Desired C |
| --- | --- | --- | --- |
| Assess a patients' readiness to engage in weight management. | 4.40 (0.76) | 4.54 (0.78) | 4.75 (0.52) |
| Communicate the benefits to overall health of sustained weight loss to persons with obesity (PwO). | 4.51 (0.76) | 4.63 (0.83) | 4.82 (0.48) |
| Develop a comprehensive, personalized obesity management care plan taking into account the patient's unique circumstances | 4.37 (0.81) | 4.41 (0.98) | 4.64 (0.62) |
| Elicit a comprehensive obesity-focused medical history. (Weight trends, previous weight loss attempts, etc.) | 4.34 (0.85) | 4.30 (1.05) | 4.64 (0.56) |
| Engage the patients in shared decision-making by incorporating their values and preferences. | 4.41 (0.79) | 4.57 (0.86) | 4.79 (0.57) |
| Perform and document a comprehensive physical examination for the assessment of obesity and the patient's overall health. | 4.45 (0.76) | 4.41 (0.96) | 4.82 (0.48) |
| Provide on-going care including the assessment of patient progress (weight, energy, function, effect on obesity related comorbidities) and supporting maintenance. | 4.39 (0.83) | 4.37 (1.06) | 4.75 (0.59) |
| Recommend (or refer for) personal and achievable behavioral interventions | 4.42 (0.76) | 4.57 (0.72) | 4.68 (0.61) |
| Recommend (or refer for) personal and achievable nutrition interventions | 4.47 (0.77) | 4.70 (0.70) | 4.79 (0.50) |
| Recommend (or refer for) personal and achievable physical activity interventions | 4.51 (0.73) | 4.63 (0.68) | 4.75 (0.59) |
| Utilize pharmacological treatments when indicated | 4.11 (1.11) | 4.48 (1.01) | 4.50 (0.88) |
| Refer appropriate patients for surgical treatments | 4.35 (0.88) | 4.67 (0.60) | 4.57 (0.74) |

Values are mean (SD). Likert scale: 1 = low agreement; 5 = high agreement.

## References

1. Wilding J. Once-Weekly Semaglutide in Adults with Overweight or Obesity. N Engl J Med. 2021 Mar 17;384(11):989–1002. doi:10.1056/NEJMoa2032183

2. Kaplan LM, Golden A, Jinnett K, Kolotkin RL, Kyle TK, Look M, et al. Perceptions of Barriers to Effective Obesity Care: Results from the National ACTION Study: Perceptions of Barriers to Effective Obesity Care. Obesity. 2018 Jan;26(1):61–9. doi:10.1002/oby.22054

3. Bagley P. Medications to Promote Weight Loss: Guidelines From the American Gastroenterological Association. Am Fam Physician. 2023 Oct;108(4):424–6.

4. Grunvald E, Shah R, Hernaez R, Chandar AK, Pickett-Blakely O, Teigen LM, et al. AGA Clinical Practice Guideline on Pharmacological Interventions for Adults With Obesity. Gastroenterology. 2022 Nov;163(5):1198–225. doi:10.1053/j.gastro.2022.08.045

5. Pedersen SD, Manjoo P, Dash S, Jain A, Pearce N, Poddar M. Pharmacotherapy for obesity management in adults: 2025 clinical practice guideline update. Can Med Assoc J. 2025 Aug 11;197(27):E797–809. doi:10.1503/cmaj.250502

6. Fox RD. Facilitating learning and change in physicians: Implications for a system of continuing medical education in Europe. Clin Microbiol Infect. 1996 Mar 1;1(3):203–7. doi:10.1111/j.1469-0691.1996.tb00554.x PubMed PMID: 11866758.

7. Fox RD, Mazmanian PE, Putnam RW, editors. Changing and learning in the lives of physicians. New York: Praeger; 1989. 194 p.

8. Bodenheimer T, Ghorob A, Willard-Grace R, Grumbach K. The 10 building blocks of high-performing primary care. Ann Fam Med. 2014;12(2):166–71. doi:10.1370/afm.1616 PubMed PMID: 24615313; PubMed Central PMCID: PMC3948764.

9. Liu L, Chien A, Singer S. Enabling System Functionalities of Primary Care Practices for Team Dynamics in Transformation to Team-Based Care: A Qualitative Comparative Analysis (QCA). Healthcare. 2023 Jul 13;11(14):2018. doi:10.3390/healthcare11142018

